# A scalable EHR-based approach for phenotype discovery and variant interpretation for hereditary cancer genes

**DOI:** 10.1101/2021.03.18.21253763

**Authors:** Chenjie Zeng, Lisa A. Bastarache, Ran Tao, Eric Venner, Scott Hebbring, Justin D. Andujar, Sarah T. Bland, David R. Crosslin, Siddharth Pratap, Ayorinde Cooley, Jennifer A. Pacheco, Kurt D. Christensen, Emma Perez, Carrie L. Blout Zawatsky, Leora Witkowski, Hana Zouk, Chunhua Weng, Kathleen A. Leppig, Patrick M. A. Sleiman, Hakon Hakonarson, Marc. S. Williams, Yuan Luo, Gail P. Jarvik, Robert C. Green, Wendy K. Chung, Ali G. Gharavi, Niall J. Lennon, Heidi L. Rehm, Richard A. Gibbs, Josh F. Peterson, Dan M. Roden, Georgia L. Wiesner, Joshua C. Denny

## Abstract

Knowledge of the clinical spectrum of rare genetic disorders helps in disease management and variant pathogenicity interpretation. Leveraging electronic health record (EHR)-linked genetic testing data from the eMERGE network, we determined the associations between a set of 23 hereditary cancer genes and 3017 phenotypes in 23544 individuals. This phenome-wide association study replicated 45% (184/406) of known gene-phenotype associations (*P* = 5.1×10^−125^). Meta-analysis with an independent EHR-derived cohort of 3242 patients confirmed 14 novel associations with phenotypes in the neoplastic, genitourinary, digestive, congenital, metabolic, mental and neurologic categories. Phenotype risk scores (PheRS) based on weighted aggregations of EHR phenotypes accurately predicted variant pathogenicity for at least 50% of pathogenic variants for 8/23 genes. We generated a catalog of PheRS for 7800 variants, including 5217 variants of uncertain significance, to provide empirical evidence of potential pathogenicity. This study highlights the potential of EHR data in genomic medicine.

---

Understanding the phenotypic consequences of genomic variation is critical to genomic medicine. Uncovering gene-phenotype relationships facilitates clinical diagnoses, leads to better treatment, improves prognosis prediction, and provides insights into disease etiology and potential therapeutic targets^1,2^. The application of next generation sequencing (NGS) has markedly accelerated the discovery of novel Mendelian disease genes and has expanded our knowledge of the characteristic phenotypes associated with genetic disorders. These are epitomized by hereditary cancer genes. It has been shown that their associated phenotypes can extend beyond predisposition to cancer^3-5^. For example, developmental disorders are often found in patients with hereditary cancer syndromes^5^. However, substantial gaps in knowledge about the spectrum of phenotypes have been noted^6^, suggesting the need for infrastructure and resources to systematically assess gene-phenotype associations^6,7^. In this study, we used electronic health record (EHR) data to systematically evaluate a wide range of phenotypes associated with hereditary cancer genes.

The clinical consequences of a genetic variant depend on the variant’s pathogenicity and its penetrance. The American College of Medical Genetics and Genomics (ACMG) has defined a set of 59 genes, including 25 associated with cancer syndromes^8^, in which variants are known to cause disorders with clearly defined phenotypes that are clinically actionable. However, our ability to predict the pathogenicity of rare genetic variants remains poor, and these actionable genes still contain many variants of uncertain significance (VUS). We have previously demonstrated that aggregating related EHR phenotypes for Mendelian diseases could aid in variant interpretation^9,10^.

Here, we used EHR and genetic testing data from 10 clinical sites in the Electronic Medical Records and Genomics (eMERGE) Network^11^ to study a broad range of phenotypes associated with hereditary cancer genes. We replicated known gene-phenotype associations in a phenome wide association study (PheWAS). We next identified new associations and replicated them in an independent cohort of patients undergoing clinical genetic testing. We then tested the utility of EHR phenotypes in assessing the pathogenicity of rare variants through the application of phenotype risk scores (PheRS)^9,10^ to aid in future variant interpretation in clinical genetic testing.

## Results

Figure 1 provides an overview of the study design. Our primary study population included 23,544 individuals from 10 sites who were sequenced on a custom NGS panel that includes 31 genes with known associations with hereditary cancers including, 25 of which were included drawn from the ACMG59 list^8^ and 6 additional genes (*ATM, BLM, CHEK2, PALB2, POLE*, and *POLD1*) selected by participating sites^11^. We assembled an independent cohort of 3242 individuals by linking the Vanderbilt hereditary cancer registry (HCR) that documented testing results of patients undergoing clinical genetic testing for hereditary cancer syndromes to the EHR database at Vanderbilt. Supplemental Table S1 summarizes the distribution of demographics and mean follow-up time for each site in the eMERGEseq cohort and the HCR cohort. The classification of variants in both cohorts was performed by the Clinical Laboratory Improvement Amendments (CLIA) and the College of American Pathologists (CAP)-accredited molecular genetic laboratories as described in the Methods section and elsewhere^11^. For each gene, we defined individuals with pathogenic/likely pathogenic (P/LP) variants as carriers and those with benign/likely benign (B/LB) variants or no rare variants (minor allele frequency < 0.001) as non-carriers. We identified 892 carriers for 23 genes in the eMERGEseq cohort. The HCR cohort included 434 carriers for 19 of these 23 genes. Distributions of carriers, non-carriers and individuals with variants of uncertain significance (VUS) for each gene are presented in Supplemental Table S2.

**Figure 1.**
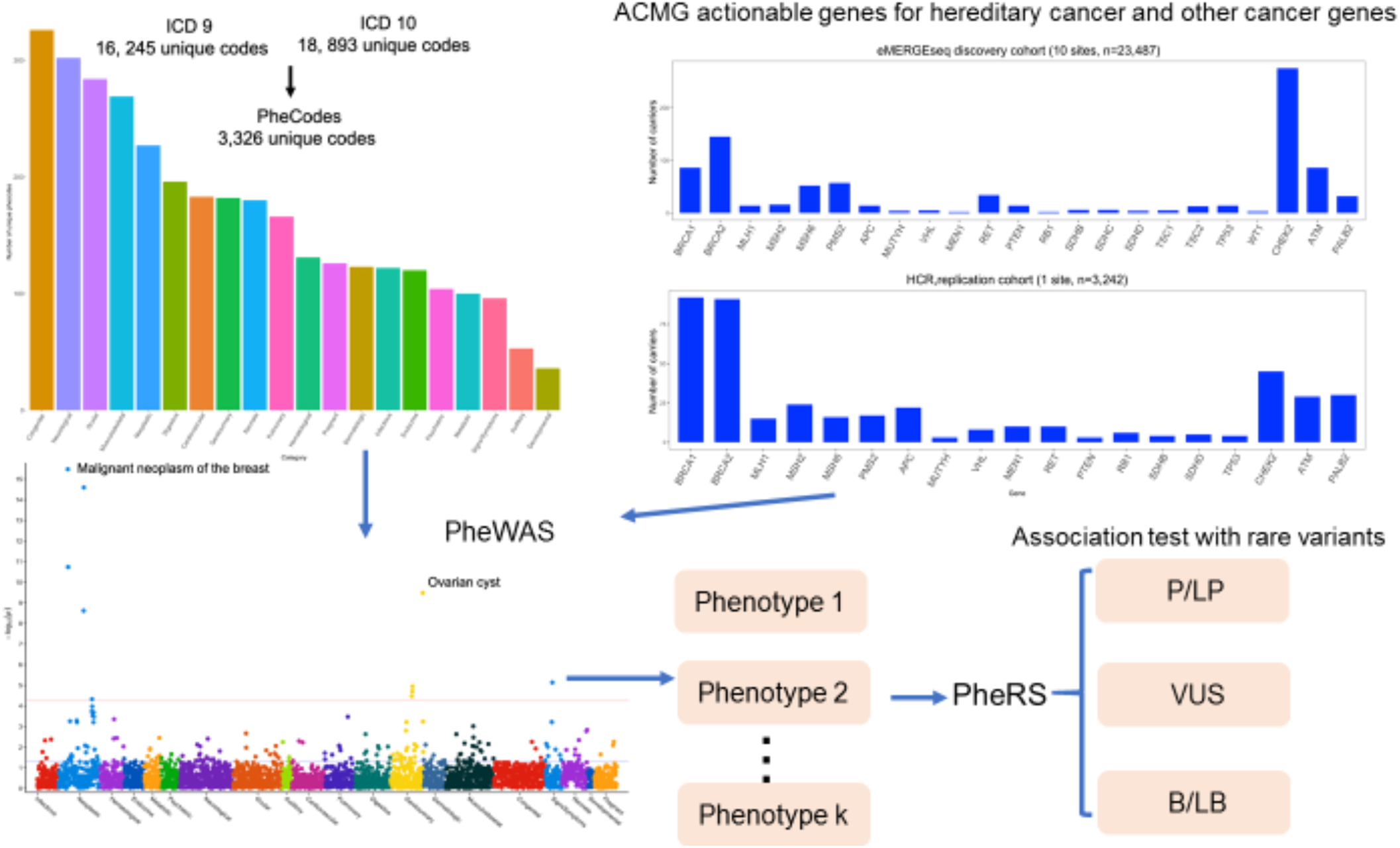
Study Flowchart. To identify phenotypes associated with hereditary cancer genes, we revised and expanded the PheWAS mapping algorithm to derive 3326 unique phenotypes from ICD9/10 codes. Leveraging genetic testing data from the Electronic Medical Records and Genomics (eMERGE) network III, we identified carriers of P/LP alleles of cancer susceptible genes in 23544 individuals from the 10 clinical sites under the eMERGE network and compared their clinical phenotypes with those of non-carriers through PheWAS. We validated new phenotypes associated with these genes in an independent cohort of 3242 patients undertaking clinical genetic testing at Vanderbilt University Medical Center (VUMC) and enrolled in the hereditary cancer registry (HCR). All variants in both studies were classified by CLIA and CAP-accredited molecular genetic testing laboratories. To explore the utility of phenotypes, we computed phenotype risk scores (PheRS) using phenotypes documented in the OMIM databases and new phenotypes identified in this study. We tested associations between PheRSs for hereditary cancer genes and rare variants. P/LP: pathogenic/likely pathogenic; VUS: variant of uncertain significance; and B/LB: benign/likely benign.

### PheWAS replicated known associations

To validate the PheWAS approach in uncovering phenotypes associated with hereditary cancer genes, we assessed whether PheWAS could replicate known gene-phenotype relationships. PheWAS replicated 184 out of 406 (45%) known gene-phenotype associations as documented in the Online Mendelian Inheritance in Man (OMIM) database (Figure 2 and Supplemental Table S2). The probability of replicating 184 associations out of 406 tests by chance, under the null hypothesis of no association, is 5.1×10^−125^. When limiting analyses to cancer phenotypes in the gene-disease relationship with definite clinical validity assessed by ClinGen^12,13^, PheWAS replicated 73% (33/45, *P* =1.8 ×10^−33^, under the null hypothesis of no association) of these gene-cancer associations (Supplemental Table S3). Thirty-two out of 38 (88%) associations with high penetrance and 44 out of 60 (73%) associations with high to moderate penetrance were replicated while only 3 out of 14 (21%) with moderate penetrance and 2 out of 11 (18%) with low penetrance were replicated (Figure 2). The most common categories of phenotypes replicated were developmental, neurological, congenital, and neoplastic.

**Figure 2.**
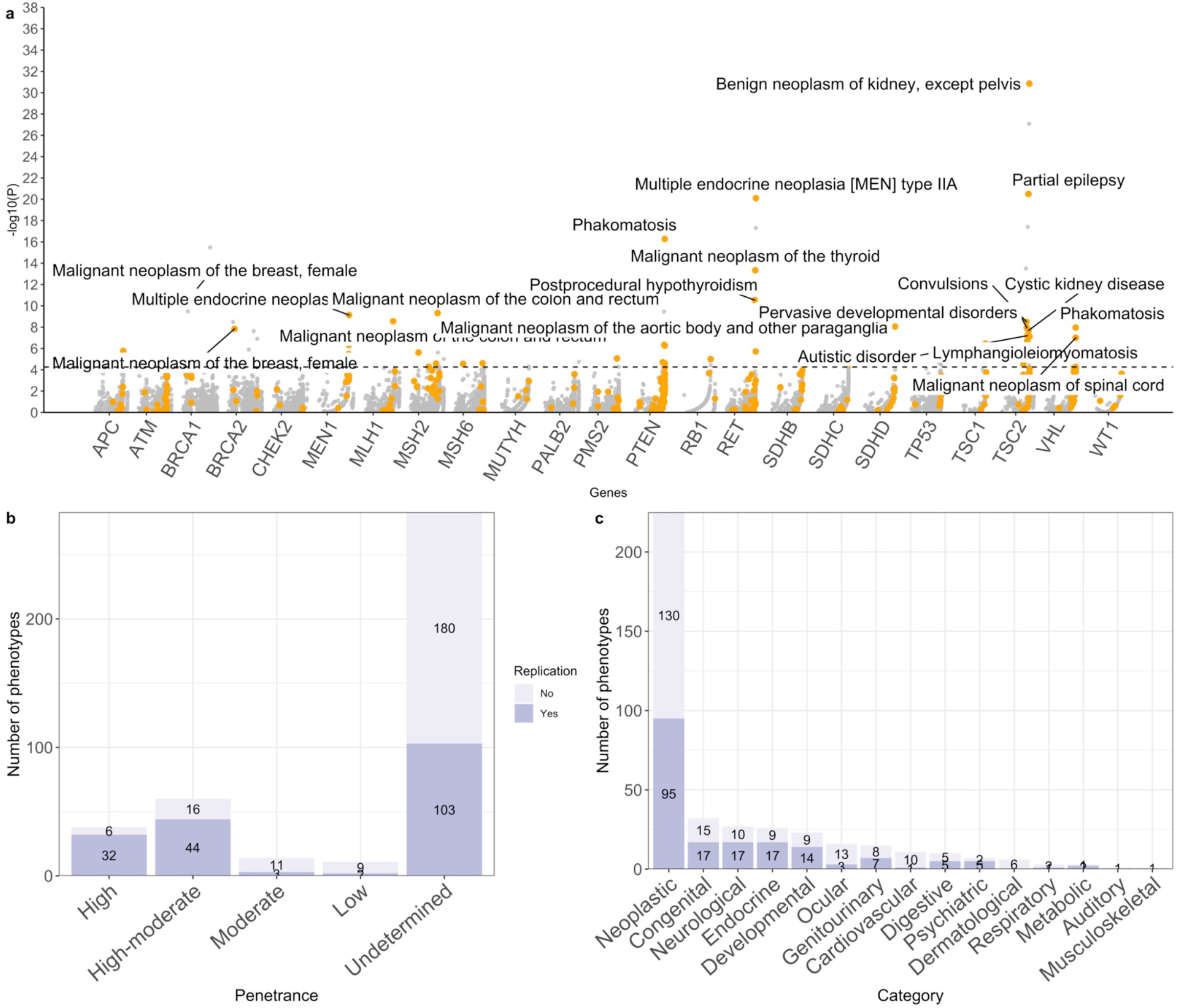
PheWAS study of 23,544 eMERGEseq participants confirms known gene-phenotype associations for cancer susceptibility genes. a. PheWAS replicated known gene-phenotype associations in the eMERGEseq cohort. Strength of the association is plotted along the y axis as -log10(P), and phenotypes are represented on the x axis, grouped by each gene. Orange dots represent the known associated phenotypes. Those with a *P* < 1× 10^−7^ are heighted. The dashed line indicates *P* = 2.5 × 10^−5^, representing the empirical phenome-wide significance for each gene. b. Number of replicated gene-phenotype associations according to penetrance of genes in the phenotypes. We defined replicated associations as associations with a *P* < 0.05 and a consistent direction of effect in the PheWAS. Penetrance categorizations based on the estimated lifetime risk are provided in Supplemental Table 3. c. Number of replicated gene-phenotype associations according to the categories of phenotypes.

A total of 42 known gene-phenotype associations exceeded the phenome-wide significance level with a *P*< 2.5×10^−5^. These associations included *BRCA1* and *BRCA2* with breast cancer, *MLH1, MSH2, MSH6*, and *PMS2* with colorectal cancer, *MSH2* with endometrial cancer, *RB1* with malignant neoplasm of the retina, *RET* with thyroid cancer, *SDHD* with paraganglioma, *TSC2* with benign neoplasm of the kidney and *PTEN* and *VHL* with phakomatosis (odds ratios (ORs) ranged from 4.8-7598.0, Supplemental Table S4).

### PheWAS identified new gene-phenotype associations

A total of 95 associations found in the eMERGEseq cohort exceeded the phenome-wide significance level at a *P* < 2.5 × 10^−5^, and 211 associations showed suggestive statistical evidence with a *P* < 5 × 10^−4^. After removing known associations and associations related to known phenotypes, six novel associations with a *P* < 2.5 × 10^−5^ were identified, namely, *BRCA1* and *BRCA2* with ovarian cysts (OR = 5.9, *P* = 3.3 × 10^−10^ and OR = 4.1, *P* = 3.3 × 10^−9^, respectively), *SDHx* (*SDHB, SDHC* and *SDHD*) with Budd-Chiari syndrome (OR = 364.5, *P* = 1.2 × 10^−6^), *TSC2* with dementia (OR = 54.9, *P* = 8.0 × 10^−6^), *MSH6* with premature separation of placenta (OR= 73.7, *P* = 9.8 × 10^−6^), and *PMS2* with other infection during labor (OR=155.1, *P* = 1.8 × 10^−5^). We also found 64 new gene-phenotype associations with suggestive evidence with a *P* < 5 × 10^−4^. Notably, among genes for the Lynch syndrome, we found evidence of association with digestive diseases, including *MLH1* with ulceration of the lower gastrointestinal (GI) tract (OR= 26.8, *P* = 8.3× 10^−5^) and *MSH6* with gastrointestinal angiodysplasia (OR = 15.0, *P* = 8.3× 10^−5^, Supplemental Table S5).

To replicate associations found in the eMERGEseq cohort and to identify additional new associations, we evaluated 6433 associations in the HCR dataset (n = 3242). Combining results from both datasets, we replicated associations of *BRCA1* and *BRCA2* with ovarian cysts, *MLH1* with ulceration of the lower GI tract, *APC* with benign neoplasms of the liver and intrahepatic bile ducts, *CHEK2* with leukemia, *PMS2* with spermatocele, and *RET* with diplopia. We also identified additional new associations, including *MSH6* with bladder cancer, *APC* with gastritis and duodenitis, *MEN1* with acute pancreatitis, *VHL* with congenital malformations of the spleen, *BRCA1* with vitamin D deficiency, *MUTYH* with polycystic ovarian syndrome, and *PMS2* with cannabis dependence. All results are presented in Table 1, Figure 3 and Supplemental Table S6.

**Table 1.** Novel associations discovered via PheWAS. All results with a *P* < 2.5 × 10^−5^ and in both cohorts with a consistent direction of effect are included.

| Gene | Phenotypes | eMERGEseq |  | HCR |  | Meta |  |
| --- | --- | --- | --- | --- | --- | --- | --- |
|  |  | OR (95% CI) | *P* | OR (95% CI) | *P* | OR (95% CI) | *P* |
| *BRCA2* | Ovarian cyst | 4.1 (2.6-6.5) | $3.3 \times 10^{-9}$ | 2.6 (1.6-4.5) | $2.9 \times 10^{-4}$ | 3.4 (2.4-4.8) | $8.3 \times 10^{-12}$ |
| *BRCA1* | Ovarian cyst | 5.9 (3.4-10.3) | $3.3 \times 10^{-10}$ | 1.8 (1.1-3.1) | 0.03 | 3.2 (2.2-4.7) | $3.4 \times 10^{-9}$ |
| *MSH6\** | Malignant neoplasm of the bladder | 8.3 (2.3-29.5) | 0.001 | 19.0 (4.3-83.3) | $9.6 \times 10^{-5}$ | 11.8 (4.5-30.9) | $5.2 \times 10^{-7}$ |
| *APC* | Benign neoplasm of the liver and intrahepatic bile ducts | 61.0 (7.7-486.0) | $1.0 \times 10^{-4}$ | 26.5 (3.5-202.3) | 0.002 | 39.8 (9.3-169.8) | $6.5 \times 10^{-7}$ |
| *APC* | Gastritis and duodenitis | 3.3 (1.0-11.3) | 0.05 | 9.4 (3.7-24.3) | $3.4 \times 10^{-6}$ | 6.5 (3.0-13.5) | $1.2 \times 10^{-6}$ |
| *MLH1* | Ulceration of the lower GI tract | 26.8 (5.1-113.9) | $9.3 \times 10^{-5}$ | 12.4 (2.0-77.5) | 0.007 | 19.0 (5.6-64.7) | $2.5 \times 10^{-6}$ |
| *MEN1* | Acute pancreatitis | 48.5 (3.1-765.5) | 0.006 | 27.3 (4.7-158.7) | $2.4 \times 10^{-4}$ | 32.2 (7.3-142.1) | $4.6 \times 10^{-6}$ |
| *CHEK2* | Leukemia | 4.4 (2.2-8.9) | $3.6 \times 10^{-5}$ | 5.0 (2.4-8.6) | 0.05 | 4.5 (2.4-8.6) | $4.9 \times 10^{-6}$ |
| *VHL* | Congenital malformations of spleen | 111.4 (6.6-1880.1) | 0.001 | 170.5 (6.1-4766.6) | 0.002 | 133.1 (15.4-1148.4) | $8.6 \times 10^{-6}$ |
| *PMS2* | Spermatocele | 20.5 (4.1-101.2) | $2.1 \times 10^{-4}$ | 19.1 (1.5-242.8) | 0.02 | 20.1 (5.2-77.7) | $1.4 \times 10^{-5}$ |
| *BRCA1* | Vitamin D deficiency | 0.5 (0.3-0.9) | 0.03 | 0.2 (0.1-0.4) | $1.6 \times 10^{-5}$ | 0.3 (0.2-0.6) | $1.4 \times 10^{-5}$ |
| *MUTYH* | Polycystic ovarian syndrome | 33.9 (2.3-501.3) | 0.01 | 53.8 (5.8-502.1) | $4.7 \times 10^{-4}$ | 44.6 (8.0-248.7) | $1.5 \times 10^{-5}$ |
| *PMS2* | Cannabis dependence | 15.7 (2.6-95.8) | 0.003 | 184.3 (8.3-4085.9) | $9.7 \times 10^{-4}$ | 29.3 (6.2-140.0) | $2.2 \times 10^{-5}$ |
| *RET* | Diplopia | 9.9 (3.0-32.2) | $1.4 \times 10^{-4}$ | 8.4 (0.9-82.7) | 0.07 | 9.6 (3.4-27.3) | $2.4 \times 10^{-5}$ |

\*Although transitional cell carcinoma has been reported with Lynch syndrome, *MSH6*'s association with this phenotype has not been clearly shown.

**Figure 3.**
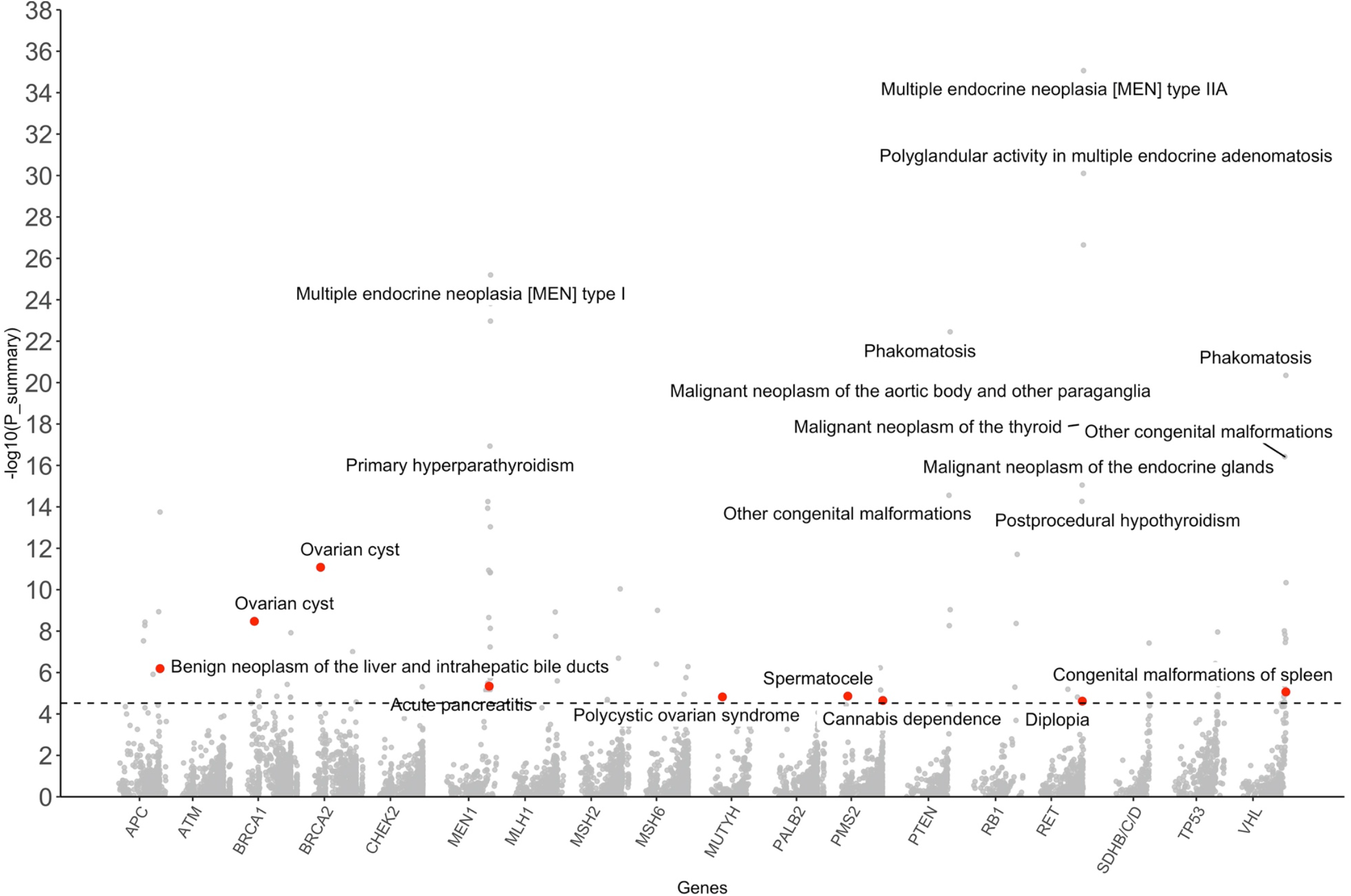
Meta-analysis of PheWAS results from the eMERGEseq and the hereditary cancer registry (HCR) at VUMC. Labeled phenotypes with red dots represent those not in OMIM that were significant in the meta-analysis of results from the eMERGEseq and the independent cohort (hereditary cancer registry at VUMC) with a *P<* 2.5 × 10^−5^. We combined *SDHB, SDHC* and *SDHD* genes into one group to increase power. *TSC1*, T*SC2*, and *WT1* are not reported as there were no carriers in the HCR set. Strength of the association estimated from the meta-analysis of results from both studies is plotted along the y axis as -log10(P_summary), and phenotypes are represented on the x axis, grouped by each gene or pathway. Known phenotypes that were highly statistically significantly with cancer genes in the meta-analysis are labeled. The dashed line indicates *P* = 2.5 × 10^−5^, representing the phenome-wide significance for each gene/pathway.

We performed conditional analyses to test for statistical independence of the novel associations from known associations (Supplemental Table S7). In the eMERGEseq cohort, after controlling for known phenotypes, all new associations remained materially unchanged except the association of *MEN1* with acute pancreatitis and *BRCA1* with vitamin D deficiency (*P* > 0.1 after adjustment of known phenotypes). In the HCR cohort, after controlling for known phenotypes, associations of *BRCA1* with ovarian cysts, *VHL* with congenital malformations of the spleen, *RET* with diplopia, and *MEN1* with acute pancreatitis attenuated with a *P* > 0.1 while the other new associations remained largely unchanged.

We conducted clinical chart reviews for all patients with readily accessible EHRs in the Synthetic Derivative (SD) and Research Derivative (RD) at VUMC to gather more information about the diagnoses related to new associations revealed in the meta-analysis. We confirmed the presence of the diagnoses by reviewing pathology reports, radiology imaging, and clinical narratives. We found patterns of co-occurrence of both novel and previously known phenotypes among the HCR patients. These included ovarian cysts with ovarian cancer in *BRCA1* carriers, diplopia with thyroid cancer in *RET* carriers, and acute pancreatitis with *MEN1* diagnoses, which might explain the attenuated associations after controlling for known phenotypes in the HCR cohort (Supplemental Table S8). Notably, we did not find pancreatic cancer diagnoses among *MEN1* carriers with acute pancreatitis. We also did not find evidence that patient with ovarian cysts were actually cases of ovarian cancer that had been misdiagnosed in *BRCA2* carriers. Approximately 50% of the ovarian cyst cases in *BRCA2* carriers were diagnosed before their genetic diagnoses, for whom findings from examinations were incidental. The remaining patients were diagnosed during screening after their genetic diagnosis, suggesting that increased screening in this population could contribute to the observed association.

This meta-analysis also revealed 21 associations with suggestive evidence with a *P* < 5 × 10^−4^ (Supplemental Table S6), including *SDHx* genes with trigeminal nerve disorders (OR = 16.0, *P* = 4.0 ×10^− 5^), *MSH2* and *MSH6* with endometrial hyperplasia (OR = 14.0, *P* = 7.3 ×10^−5^ and OR = 10.7, *P* = 5.7 ×10^− 5^, respectively), *BRCA1* with leiomyoma of uterus (OR= 2.3, *P* = 1.4 ×10^−4^), *PMS2* with infections of genitourinary tract in pregnancy and intrauterine death (OR =10.8, *P* = 2.3 ×10^−4^ and OR= 32.0, *P* = 2.4 ×10^−5^, respectively), and *MSH6* with rupture of uterus (OR=16.7, *P* = 3.9 ×10^−4^).

We also evaluated carriers with variants with markedly reduced penetrance. It remains unclear whether *MUTYH* heterozygotes in the general population were at a higher risk of colon cancer or polyps^14^. To study the impact of *MUTYH* heterozygous variants, we conducted a PheWAS in the eMERGEseq population of individuals with one P or LP variant only. The most statistically significant associations were found with cystitis (OR = 2.1, *P* = 9.1 ×10^−7^) and sinusitis (OR = 1.5, *P* = 8.6 ×10^−6^). No evidence of association with colorectal cancer or polyps was found (Supplemental Figure 3). For the 188 *APC* I1370K carriers, the most statistically significant associations were found with noise effects on the inner ear (OR = 45.7, *P* = 6.6 ×10^−6^), disorders of refraction and accommodation (OR = 0.4, *P* = 1.5 ×10^− 5^) as well as benign prostatic hyperplasia (OR= 3.1, *P* = 3.0 ×10^−5^, Supplemental Figure 4).

### PheRS provided evidence for the pathogenicity of rare variants

To test whether PheRSs that aggregate EHR phenotypes could aid in assessing the pathogenicity of variants, we first derived PheRS based on phenotypes curated by OMIM as previously described^9,10^ and explored their associations with 7800 rare variants of the 23 genes, including 377 classified as P/LP, 1 as risk allele, 2205 as B/LB, and 5217 as VUS in the eMERGEseq cohort. We tested associations between PheRSs and these variants using linear regression, assuming a dominant genetic model for variants in all genes except those in *MUTYH* for which a recessive model was assumed. Regression coefficients (betas) and *P* values are presented in Supplemental Table S9. Using the criteria of a positive beta estimate and a nominal *P* < 0.05 for each association test, 71 out of 377 P/LP variants (18%) showed evidence of pathogenicity, while only 65 out of 2205 B/LB variants (3%) showed evidence of pathogenicity (under the null hypothesis of no association, *P* = 1.2 ×10^−26^). With the same criteria, 181 out of 5217 (3%) VUS showed evidence of pathogenicity. When evaluating by each gene separately, at least 50% of the P/LP variants in genes *MEN1* (1/1*), MUTYH* (1/1), *PTEN* (6/9*), RB1*(1/2), *SDHD* (1/2), *TSC1* (2/4), *TSC2* (10/10) and *VHL* (3/4) showed evidence of pathogenicity.

To test the potential utility of new phenotypes identified in the PheWAS to improve PheRS for predicting pathogenicity of untested variants, we derived new PheRSs for genes with new phenotypes by incorporating these phenotypes in addition to OMIM phenotypes. Regression coefficients and *P* values of the new PheRSs with rare variants are presented in Supplemental Table S10. Results with a *P* < 0.001 are showed in Table 2. Distributions of regression coefficients of variants of each category of interpretation by each gene are presented in Figure 4. As expected, a larger beta for P/LP variants in the new PheRS compared with OMIM PheRS for all genes evaluated were observed. For VUS, which were not analyzed in the PheWAS analyses, a wider distribution of betas was observed for genes *APC, RET, TSC2* and *VHL* than that of B/LB variants while no such differences were found for *BRCA1/2, PMS2, MLH1* and *MSH6*. The PheRS for 188 VUS were associated with an increased risk of diseases, while another 164 VUS were associated with a reduced risk, suggesting a protective effect (Supplemental Table S10). For example, we found that a VUS variant *APC* c.385G>C was associated with an elevated PheRS for familial adenomatous polyposis (FAP) (beta = 2.3, *P* = 5.6 × 10^−6^). Clinical profiles of carriers of this variant suggested an attenuated form of FAP. Notably, 2 out of 4 patients were also with the diagnosis of gastritis and duodenitis, a new phenotype identified in the PheWAS.

**Figure 4.**
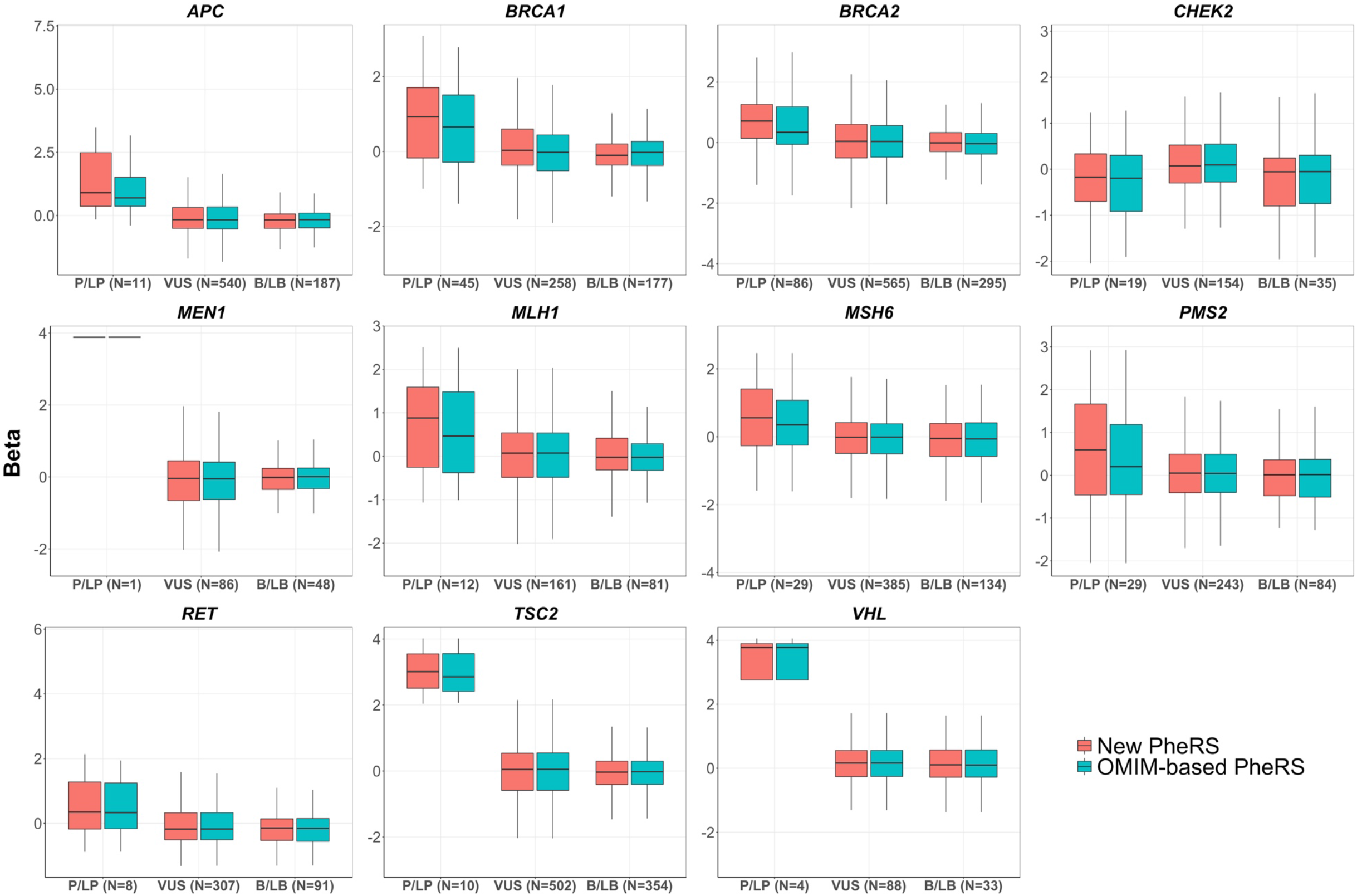
Phenotype risk scores provides evidence for the assessment of the pathogenicity of rare variants. Regression coefficients for new PheRS and OMIM PheRS on rare variants. We derived new PheRS by incorporating new phenotypes. The detailed information of the new phenotypes is presented in Supplemental Table 11. Y axis represents the beta coefficients for both PheRSs for each variant. The x axis represents the classification for the variants.

**Table 2.** Association results of PheRS for hereditary cancer syndromes. List includes all rare variants with *P* < 0.001 in the eMERGEseq cohort. Positive betas are associated with an increased risk of disease, and negative betas are associated with a decreased risk of disease.

| Gene | Variant | Diseases | No. of HET | Variant Interpretation | OMIM PheRS |  | New PheRS |  |
| --- | --- | --- | --- | --- | --- | --- | --- | --- |
|  |  |  |  |  | Beta | P* | Beta | P* |
| *RET* | c.2671T>G (p.Ser891Ala) | Multiple endocrine neoplasia II | 13 | P | 1.9 | $2.4 \times 10^{-12}$ | 2.1 | $1.2 \times 10^{-14}$ |
| *APC* | c.4594G>A (p.Asp1632Asn) | Familial adenomatous polyposis | 1 | VUS | 7.0 | $1.8 \times 10^{-12}$ | 7.1 | $1.2 \times 10^{-14}$ |
| *APC* | c.694C>T (p.Arg232Ter) | Familial adenomatous polyposis | 1 | P | 4.4 | $1.1 \times 10^{-5}$ | 6.1 | $8.6 \times 10^{-10}$ |
| *RET* | c.1858T>C (p.Cys620Arg) | Multiple endocrine neoplasia II | 1 | P | 5.7 | $1.1 \times 10^{-8}$ | 5.6 | $1.7 \times 10^{-8}$ |
| *MEN1* | c.307delC (p.Leu103fs) | Multiple endocrine neoplasia I | 2 | P | 3.9 | $3.7 \times 10^{-8}$ | 3.9 | $3.7 \times 10^{-8}$ |
| *TSC2* | c.1792T>C (p.Tyr598His) | Tuberous sclerosis | 3 | P | 3.0 | $1.8 \times 10^{-7}$ | 3.1 | $9.6 \times 10^{-8}$ |
| *RET* | c.626-4G>A** | Multiple endocrine neoplasia II | 3 | VUS | 3.1 | $7.2 \times 10^{-8}$ | 3.1 | $1.2 \times 10^{-7}$ |
| *APC* | c.385G>C (p.Glu129Gln) | Familial adenomatous polyposis | 4 | VUS | 2.2 | $1.2 \times 10^{-5}$ | 2.3 | $5.6 \times 10^{-6}$ |
| *RET* | c.431G>A (p.Arg144His) | Multiple endocrine neoplasia II | 1 | VUS | 4.6 | $4.3 \times 10^{-6}$ | 4.5 | $6.4 \times 10^{-6}$ |
| *BRCA1* | c.68_69delAG (p.Glu23Valfs) | Hereditary breast and ovarian cancer | 15 | P | 0.8 | $1.3 \times 10^{-3}$ | 1.1 | $2.3 \times 10^{-5}$ |
| *TSC2* | c.5347G>A (p.Glu1783Lys) | Tuberous sclerosis | 2 | VUS | -2.9 | $4.5 \times 10^{-5}$ | -2.9 | $4.8 \times 10^{-5}$ |
| *VHL* | c.500G>A (p.Arg167Gln) | Von Hippel-Lindau disease | 1 | P | 4.1 | $4.9 \times 10^{-5}$ | 4.1 | $4.9 \times 10^{-5}$ |
| *TSC2* | c.4495_4512del (p.1502_arg1507del) | Tuberous sclerosis | 1 | P | 4.0 | $5.7 \times 10^{-5}$ | 4.0 | $5.7 \times 10^{-5}$ |
| *TSC2* | c.3750C>G (p.Tyr1250Ter) | Tuberous sclerosis | 1 | P | 3.9 | $9.0 \times 10^{-5}$ | 3.9 | $9.2 \times 10^{-5}$ |
| *VHL* | c.432_438del (p.Glu145Trpfs) | Von Hippel-Lindau disease | 1 | P | 3.8 | $1.2 \times 10^{-4}$ | 3.8 | $1.2 \times 10^{-4}$ |
| *APC* | c.2600C>T (p.Thr867Ile) | Familial adenomatous polyposis | 1 | VUS | 3.6 | $3.6 \times 10^{-5}$ | 3.8 | $1.7 \times 10^{-4}$ |
| *APC* | c.5241G>A (p.Met1747Ile) | Familial adenomatous polyposis | 1 | VUS | 4.1 | $4.4 \times 10^{-4}$ | 3.7 | $1.8 \times 10^{-4}$ |
| *VHL* | c.264G>T (p.Trp88Cis) | Von Hippel-Lindau disease | 1 | LP | 3.7 | $2.1 \times 10^{-4}$ | 3.7 | $2.1 \times 10^{-4}$ |
| *TSC2* | c.4581dup (p.Glu1528Ter) | Tuberous sclerosis | 1 | P | 3.7 | $2.3 \times 10^{-4}$ | 3.7 | $2.3 \times 10^{-4}$ |
| *BRCA2* | c.3974_3975insTGCT (p. Ala1325Cysfs) | Hereditary breast and ovarian cancer | 1 | P | 3.3 | $8.7 \times 10^{-4}$ | 3.5 | $4.0 \times 10^{-4}$ |
| *APC* | c.1409-2A>G** | Familial adenomatous polyposis | 1 | LP | 3.2 | $1.6 \times 10^{-3}$ | 3.5 | $4.8 \times 10^{-4}$ |
| *TSC2* | c.1070C>T (p.Ala357Val) | Tuberous sclerosis | 21 | VUS | 0.8 | $5.2 \times 10^{-3}$ | 0.7 | $6.4 \times 10^{-4}$ |
| *BRCA2* | c.7068_7069del (p.Gln2356Hisfs) | Hereditary breast and ovarian cancer | 4 | P | 1.6 | $1.2 \times 10^{-3}$ | 1.7 | $7.8 \times 10^{-4}$ |

\**P* values for singletons are likely not stable, although betas for these variants could represent evidence of possible pathogenicity. \*\* *RET* c.626-4G>A is an intron variant, *APC* c.1409-2A>G is a splice acceptor variant.

## Discussion

We present a scalable approach to discover new phenotypes related to hereditary cancer genes and evaluate the pathogenicity of variants using EHR data. We demonstrated the validity of the PheWAS approach by replicating 73% of the established primary gene-cancer associations and 45% of all gene-phenotype associations documented in OMIM. PheWAS also revealed new phenotypes that were replicated in an independent EHR-derived cohort. PheRS that aggregated associated phenotypes predicted pathogenicity of rare variants and provided evidence for the pathogenicity of 5217 VUS in hereditary cancer genes.

This study demonstrated the feasibility of rapid phenotype discovery of rare genetic disorders in EHR data. PheWAS replicated nearly half of the gene-phenotype relationships documented in the OMIM database that curates many decades of knowledge of these genes^15^. Replication rates were significantly higher among those with a higher estimated life-time risk, supporting that EHR data could recapitulate previous findings in a relatively precise manner. Many of the nonreplicated associations were related to disorders or symptoms which were more likely to be under-documented in the system of billing codes, such as dental caries and pigmentation disorders. Future studies that incorporate additional data such as clinical notes and images will improve the resolving power.

This study revealed 14 novel gene-phenotype associations, for which phenotypes were found in the genitourinary, digestive, congenital, metabolic, mental and neurologic categories in addition to neoplasms. These findings, although yet to be validated in additional studies, further support that hereditary cancer syndromes can have a broad clinical spectrum^3^. Most of these associations would have been difficult to detect in observational studies that are primarily designed based upon prior knowledge^16^. For example, this analysis revealed associations of *PMS2* with spermatoceles and cannabis dependance that are not typically on the radar for Lynch syndrome. Chart review results suggested that some of the phenotypes could be symptoms of underlying diseases that had been known. For example, the association of *RET* with diplopia was likely to be mediated by neuroendocrine disorders including tumors. However, diplopia has been largely underreported in MEN2 patients in previous studies and thus has not been documented in the OMIM database. We believe that it is important to recognize relevant symptoms in the EHR systems, which can serve an early sign of underlying diseases such as cancers and thus facilitate early detection. Similarly, we also reported a suggestive association of *SDHx* genes with trigeminal nerve disorders that could be an early sign of paragangliomas. We also observed that increased screening in carriers could contribute to some associations, including *BRCA1/2* and ovarian cyst and *VHL* with congenital malformations of spleen. Although results from chart reviews suggested that a remarkable proportion of these patients were diagnosed with indications other than screening, additional studies are needed to elucidate these associations.

Our results generate new data for hypotheses around the pathogenesis of some common diseases. We found that *MEN1* carriers were associated with a 32-fold increased risk of acute pancreatitis. This was consistent with a recent study that identified an essential role of *MEN1* in exocrine pancreas homeostasis in response to inflammation that contributes to pancreatitis in mouse models^17^. Several previous studies suggested that *MUTYH* contributed to inflammatory-related disorders^18^. We found that *MUTYH* homozygotes or compound heterozygotes were associated with polycystic ovarian syndrome, for which chronic inflammation has been proposed to be a key contributor^19^. We also found a Bechet’s syndrome diagnosis in a *MUTYH* compound heterozygote. Additionally, we found that *MUTYH* heterozygotes were more likely to develop cystitis and sinusitis compared to non-carriers. These together provide supporting evidence for a role of *MUTYH* in disorders with an inflammatory basis.

Rapid and accurate variant interpretation remains a challenge in clinical genetics. We have previously showed that PheRS could augment variant interpretation^9,10^. Using PheRS including new phenotypes identified from the PheWAS serves as a test of the potential utility of these new phenotypes. As expected, we observed remarkable differences in the predicted pathogenicity by PheRS between pathogenic and benign variants. We found that the majority of VUS were not associated with an elevated PheRS, which was consistent with previous studies showing that the majority of VUS would be downgraded to benign if reclassified^20^. Nevertheless, we found that by adding new phenotypes into the PheRS, several VUS were associated with a higher predicted level of pathogenicity (a larger beta), suggesting that the possibility that employing additional related phenotypes could improve variant interpretation. Replication in additional studies will be needed to evaluate the pathogenicity of these VUS.

Limitations of this study include the use of phecodes, which are phenotypes based on aggregations of related billing codes. While phecodes have been shown to be an effective tool for replicating genetic associations with EHR data^21-23^, they are unable to capture all phenotypes, including some unique characteristics of hereditary cancer syndromes. For example, patients with familial adenomatous polyposis typically present with numerous polyps, a condition which lacks a specific billing code. This analysis did not take into account the specific age of disease diagnosis, adjusting for the last age of the participant documented instead. Our next step is to develop algorithms for deep phenotyping to identify detailed characteristics of cancers and other diseases through analyses of images, laboratory measurements and clinical narratives.

In summary, we demonstrated that PheWAS in EHR datasets has potential for phenotype expansion of hereditary cancer genes. We showed that aggregating clinically significant alleles increased the power to detect phenotypes, which is particularly meaningful for rare genetic disorders with smaller study cohorts. Studying rare disorders at the population level requires very large cohorts. Just as EHR-linked genotyping array data enabled the rapid expansion of GWAS cohorts for the discovery of new associations, EHR-linked sequence data will provide a similar resource to expand our knowledge of the phenotypic consequences of Mendelian disease-causing genes. We anticipate that applying these approaches to large datasets such as the UK Biobank^24^ and the *All of Us* Research Program^25^ will help reveal the true clinical spectrum of genetic diseases, aid in variant interpretation, and ultimately facilitate precision medicine.

## Methods

### Study populations

The eMERGEseq cohort is comprised of 24956 biobank or prospectively recruited individuals from ten clinical sites under the eMERGE network. These individuals were either unselected or were enriched for specific clinical phenotypes depending on site-specific interest^11^. For cancer-related phenotypes, the UW/KPW site was enriched for individuals with colorectal cancer/polyps diagnoses. Additionally, two sites, CCHMC and CHOP, included pediatric patients. The major goal of this project was to study and improve the process of returning actionable genetic results to clinicians and patients^26^. A detailed description of each site, including enrollment criteria, specific research interest and enrichment of phenotypes, was reported elsewhere^11^. All studies were approved by local Institutional Review Boards (IRBs). For this study, we removed individuals without International Classification of Diseases (ICD) codes in the EHRs. A total of 23544 individuals were retained for analyses.

The replication dataset was obtained from the hereditary cancer registry (HCR) at Vanderbilt University Medical Center (VUMC), which included 3794 individuals who received clinical genetic testing for hereditary cancer from 2012 to 2020. This study was approved by the IRB at VUMC. We obtained the EHR data of 3739 individuals through the Research Derivative (RD), a database of clinical and related data derived from EHR systems^27^. Through reviewing clinical charts in the RD and records in the HCR, we removed patients who were participants of the eMERGEseq project (n=14) and family members of the index patients who were enrolled in the registry due to cascade testing (n=483). A total of 3242 patients retained for analyses. This cohort was enriched for individuals at a high risk of hereditary cancer syndromes, with 98% reporting a family history of cancers and 65% reporting a personal history of cancer. Specifically, approximately 50% of all patients reported a breast cancer diagnosis. This cohort also included pediatric cancer patients. The ages at the first cancer diagnoses ranged from 1 year old to 90 years old, with a mean age of 50.4 years old, as documented in the EHRs.

### Gene panel and sequencing in eMERGEseq and genetic testing in HCR

Details on the design of the sequencing panel of the eMERGEseq project have been previously described^11^. Briefly, this panel comprises a total of 109 genes and 1550 single nucleotide variants (SNVs). The 109 genes include 58 genes from the American College of Medical Genetics and Genomics (ACMG59) actionable finding list^8^. Additionally, each of the participating sites nominated 6 genes relevant to site-specific research interest. In this study, we focused on hereditary cancer genes on this panel, including 25 ACMG genes: *APC, BMPR1A, BRCA1, BRCA2, MEN1, MLH1, MSH2, MSH6, MUTYH, NF2, PMS2, PTEN, RB1, RET, SDHAF2, SDHB, SDHC, SDHD, SMAD4, STK11, TP53, TSC1, TSC2, VHL*, and *WT1*, and 6 cancer-related genes nominated by participating sites: *BLM, CHEK2, POLD1, POLE, PALB2*, and *ATM*.

The genetic testing in the HCR was performed by commercial molecular diagnostic laboratories.

### Classification of variants

Variant classifications in the eMERGEseq were performed by two laboratories at the sequencing centers, according to ACMG/Association of Medical Pathology (ACMG/AMP) guidelines and some specific modifications from ClinGen Sequence Variant Interpretation Working Group and ClinGen Expert Panels as previously described^11^. Variants were classified into pathogenic (P), likely pathogenic (LP), variant of uncertain significance (VUS), likely benign (LB) and benign (B). No P/LP variants for cancer syndromes were detected for *BLM, BMPR1A, NF2, POLD1, POLE, SDHAF2, SMAD4*, and *STK11*^11^. We only included variants with an allele fraction > 30%.

Variant classification in the HCR was performed by CLIA and CAP-accredited molecular genetic testing laboratories. We compared results of classification for the same variants identified in both datasets (n=11). The variant *VHL* p.R200W was classified as a VUS in the eMERGEseq dataset, while it was classified as a pathogenic variant (P) in the HCR dataset. Previous studies showed that this variant was not associated with the von Hippel Lindau (VHL) disease but congenital erythrocytosis^28^. Therefore, we considered this variant a VUS in the analysis in the HCR dataset. The interpretations for the remaining 10 variants were consistent between these two datasets.

For each gene, we defined patients with P/LP variants as carriers and patients with B/LB variants or no rare variants as non-carriers, and patients with VUS as VUS carriers. The distribution of carriers, non-carriers, and patients with VUSs for each gene is listed in Supplemental Table 2.

### PheWAS phenotyping

PheWAS phenotypes were defined using phecodes, which are manually grouped ICD-9 and ICD-10 codes developed to facilitate EHR based genetic research. In this study, we modified and expanded our previous phecode map (version 1.2) that linked ICD codes to 1967 phenotypes^29,30^ by adding more granular phenotypes, including those related to Mendelian disorders and other traits in the congenital, neonatal, developmental, ocular, and pregnancy categories. Using the EHR data from a cohort of 2.6 million patients from the Synthetic Derivative (SD) at VUMC^31^, this new algorithm derived 3368 phecodes from 16245 unique ICD-9 codes and 18893 unique ICD-10 codes, spanning the following categories: auditory, cardiovascular, congenital, dermatologic, developmental, digestive, endocrine, genitourinary, hematopoietic, infectious, musculoskeletal, neonate, neoplastic, ocular, pregnant, psychiatric, pulmonary, and symptoms/signs.

We derived 3186 unique PheWAS phenotypes from 2,134,933 unique dates of ICD-9 and -10 codes in the EHRs of the eMERGEseq cohort. We removed phenotypes with < 5 cases. A total of 3017 phenotypes remained. To empirically estimate the phenome-wide significant *P*-value threshold, we conducted 10,000 PheWAS with a random variable using data from the eMERGEseq cohort and analyzed the distributions of minimum *P*-values (Pmin) for each PheWAS. The 95^th^ percentile of Pmin was 2.5 × 10^−5^, and we defined this *P-*value as the empirical phenome-wide significance threshold at a significance level of α = 0.05 (Supplemental Figure 1), which was equivalent to the Bonferroni correction of 2000 independent tests. We also defined a suggestive association *P-*value threshold by 1 divided by the number of independent tests, which was 5 × 10^−4^ in this study, representing the level where, under the null hypothesis, one false positive is expected per phenome scan, as proposed by Lander and Kruglyak for a genome-wide scan^32^.

### Identification of known gene-phenotype associations

We retrieved the clinical synopses for each gene from the Online Mendelian Inheritance in Man (OMIM), a comprehensive, authoritative compendium of human genes and genetic phenotypes^15^, which have been annotated with the Human Phenotype Ontology (HPO)^33^. We modified some of the associations according to other authoritative resources including Orphanet^34^, GeneReviews,,^35^ and the National Comprehensive Cancer Network (NCCN) guidelines^36-41^. We also retrieved data of gene-diseases validity from ClinGen^12^. We defined gene-cancer associations with definitive evidence of clinical validity by ClinGen working groups as established gene-cancer associations^13^. We also obtained data on the life-time risk of each phenotype for carriers of each gene from these resources as well as from the most recent analyses from large cohorts^42-46^. The list of known associations was reviewed by CJZ and GLW, representing an *ad hoc* assessment of a combination of literature review and clinical expert review, to categorize associations by penetrance. The complete list of gene-phenotype associations and results of these associations using PheWAS phenotypes in the eMERGEseq cohort are found in Supplemental Table S3.

### PheWAS analyses

In the eMERGEseq cohort, we used a minimum code count threshold of one phecode to define cases for a phenotype. We defined controls as those who never had the phecode. We included genes with at least 2 carriers. We focused on phenotypes that were documented in the carriers. The number of phenotypes found in carriers for each gene is shown in Supplemental Figure 2. For phenotypes found in carriers, each gene-phenotype association was tested independently using the firth logistic regression adjusted for age, unique years of records in the EHR, sex, eMERGE sites, and the first 4 principal components (PCs). Analyses were performed assuming an autosomal dominant inheritance for all genes with the exception of *MUTYH*, for which an autosomal recessive inheritance was assumed. For phenotypes only found in the non-carriers, we performed a supplementary Fisher’s exact test analysis to evaluate their associations with the gene. To increase the power to uncover new phenotypes for genes with less than 10 carriers, we grouped genes into the same pathway according to their molecular functions and clinical spectrums if available. Specifically, we grouped *SDHB, SDHC* and *SDHD* into one pathway (“*SDHx*”).

We categorized all associations found in the eMERGEseq into three groups: known associations, associations related to known associations (elevated cancer antigen 125 for *BRCA1/2*, for example), and potentially new associations.

We considered a known phenotype-gene association replicated in our analysis if the PheWAS had a *P* < 0.05 with the expected direction of effect between phenotype and genetic variant. Using the same approach as we previously reported^30^, to test the probability of replicating X out of Y known associations at α = 0.05, we calculated based on the probability of drawing *P*-values randomly from a normal distribution with at least X of them having a *P* < 0.05 (X being the number of replicated associations). Thus, the probability of getting X gene-phenotype associations replicated (*P* < 0.05) out of Y tested associations is:*P(X)=C(Y,X)P*^*X*^*(1-P)*^*Y-X*^, where *P* = 0.05 and C(Y,X) represents the number of combinations among Y items selecting X.

### Replication in the HCR dataset

The HCR cohort is comprised of patients who received hereditary cancer panel testing and are thus at a higher risk of hereditary cancer syndromes compared with the general population, with 98% reporting a family history of cancer and 65% reporting a personal history of cancer. It is likely that carriers and non-carriers in this cohort are more similar in clinical phenotypes, compared to those representing the general population. If associations were replicated in this cohort, the probability that they were replicated in cohorts based on general populations would be higher.

In the HCR dataset, for phenotypes found in carriers, each gene-phenotype association was tested independently using firth logistic regression adjusted for age, years in records, sex, and race documented in the EHR, assuming an autosomal dominant inheritance for all genes with the exception of *MUTYH*, for which an autosomal recessive inheritance was assumed. The number of phenotypes evaluated in each gene is shown in Supplemental Figure 2. A total of 6433 gene-phenotype associations were found in both the eMERGEseq and the HCR datasets. A fixed-effect meta-analysis was used to estimate the summarized effect size and *P*-values. All *P*-values in this study were two-sided.

We performed conditional analyses to determine whether the new associations were associated with known phenotypes by adjusting for known phenotypes in the regression.

We conducted chart reviews to confirm the diagnoses of the PheWAS phenotypes and to study the relationships between new and known phenotypes. Specifically, for the PheWAS phenotype ovarian cysts, we reviewed all cases among *BRCA1/2* carriers in the HCR cohort (21 and 24 cases among carriers of *BRCA1* and *BRCA2*, respectively). We also reviewed a random subset of 54 cases among non-carriers of *BRCA1/2*. We tested the hypothesis that there was no difference in terms of the PheWAS phenotype capturing the actual diagnoses for ovarian cyst between carriers and non-carriers. Under this null hypothesis, *P* =1 for both genes. Results of clinical chart review are found in Supplemental Table 8.

### PheRS analyses

#### PheRS based on OMIM phenotypes

As previously described^9,10^, the PheRS for each gene for each individual is calculated as the sum of clinical features (phecodes) observed in a given individual weighted by the log inverse prevalence of the phecode. For genes with more than one primary diseases, we combined related diseases into one single feature set (Supplemental Table S2). For example, we combined breast cancer and pancreatic cancer for the gene *PALB2*, termed as *PALB2*-associated cancers. For phecodes that are specific to a subgroup of the population, we used the total number of this subpopulation as the denominator to calculate the prevalence. For sex-specific traits, we used the number of patients of the specific sex as the denominator. For example, for prostate cancer, we used the total number of male patients as the denominator. For phenotypes in the pregnancy category, we used the number of patients with at least one of the pregnant phenotypes in the cohort as the denominator. Weights for each phecode were calculated as the negative log inverse prevalence in the eMERGEseq cohort. For each gene and each individual, the raw PheRS was calculated by summing the weight of each phecode in the EHRs. These PheRSs were termed as the OMIM PheRSs.

#### PheRS based on OMIM and new phenotypes

We derived new PheRSs for genes *APC, BRCA1, BRCA2, CHEK2, MLH1, MSH6, MEN1, PMS2, RET, TSC2*, and *VHL* by incorporating new, significantly associated phenotypes identified in this study. The new phenotypes that were included in the new PheRS are presented in Supplemental Table S11. Similarly, we calculated the weight for each phecode as the negative log inverse prevalence in the eMERGEseq cohort. We calculated PheRSs by summing the weights of each phecode documented in the EHRs. These PheRSs were termed as the new PheRSs.

#### Assessment of pathogenicity of rare variant through PheRS

We derived residual PheRS (rPheRS) using a linear regression with the raw PheRS as the outcome, adjusted for age, sex, number of unique years in the EHR and the first 4 PCs. We applied inverse normal transformation to transform the rPheRS if the skewness was larger than 2.

We tested the association of all rare variants in the genes with the rPheRS or transformed rPheRS using a multivariate linear model adjusted for the first 4 PCs. We calculated *P* values using the score statistics. We consider associations with a *P* < 0.05 and a positive beta estimate indicating an increased burden of diseases as evidence of pathogenicity and associations with a *P* < 0.05 and a negative beta estimate indicating a decreased burden of diseases as evidence of protectiveness. Results of associations of PheRSs including the new PheRS and the OMIM PheRS with rare variants are presented in Supplemental Table S10.

## Supporting information

Supplemental Figures and part of the Supplemental Tables

## Data Availability

Genetic and phenotype data of the eMERGEseq cohort are publicly available in the dbGaP repository under phs001616.v1.p1. All summary statistics for significant gene-phenotype associations from the eMERGEseq and the HCR cohorts are provided in the Supplemental Tables S3-6. All summary statistics for associations of PheRS with genetic variants are provided in Supplemental Tables S9-10.

## Acknowledgments

Support for the research and personnel was provided by the R01LM010685 grant from the National Library of Medicine and the eMERGE grants. The eMERGE sites were funded through several series of grants from the National Human Genome Research Institute: U01HG8657, U01HG006375, U01HG004610 (Kaiser Permanente Washington/University of Washington); U01HG8685 (Brigham and Women’s Hospital); U01HG8672, U01HG006378, U01HG004608 (Vanderbilt University Medical Center); U01HG8666, U01HG006828 (Cincinnati Children’s Hospital Medical Center); U01HG6379, U01HG04599 (Mayo Clinic); U01HG8679, U01HG006382 (Geisinger Clinic); U01HG008680 (Columbia University Health Sciences); U01HG8684, U01HG006830 (Children’s Hospital of Philadelphia); U01HG8673, U01HG006388, U01HG004609 (Northwestern University); U54MD007593, U54MD007586 (Meharry Medical College); U01HG8676 (Partners Healthcare/Broad Institute); U01HG8664 (Baylor College of Medicine); U01HG006389 (Essentia Institute of Rural Health, Marshfield Clinic Research Foundation and Pennsylvania State University); U01HG006380 (Icahn School of Medicine at Mount Sinai); U01HG8701, U01HG006385, U01HG04603 (Vanderbilt University Medical Center serving as the Coordinating Center); eMERGE Genotyping Centers were also funded through U01HG004438 (CIDR) and U01HG004424 (the Broad Institute). Vanderbilt University Medical Center’s Synthetic Derivative, Research Derivative and BioVU are supported by institutional funding and by the CTSA grant ULTR000445 from NCATS/NIH. The majority of CJZ’s work on this project was supported by T32 CA160056 (NCI). The majority of JCD’s work on this project was while he was on faculty at Vanderbilt University before joining the NIH.

## Data and code availability

Genetic and phenotypic data of the eMERGEseq cohort are publicly available in the dbGaP repository under phs001616.v1.p1. All summary statistics for significant gene-phenotype associations from both the eMERGEseq and the HCR cohorts are provided in the supplemental Table S3-6. All summary statistics for associations of PheRS with genetic variants are provided in Supplemental Table S9-10. Codes for PheWAS and PheRS analyses will be available at https://github.com/chenjiezeng/CancerPheWAS.

**A scalable EHR-based approach for phenotype discovery and variant interpretation for hereditary cancer genes**

