## Supplemental Figures and part of the Supplemental Tables for "A scalable EHR-based approach for phenotype discovery and variant interpretation for hereditary cancer genes"

**Affiliations**
1. Department of Medicine, Vanderbilt University Medical Center, Nashville, TN
2. Center for Precision Medicine, Department of Biomedical Informatics, Vanderbilt University Medical Center, Nashville, TN
3. Department of Biostatistics, Vanderbilt Genetics Institute, Vanderbilt University Medical Center, Nashville, TN
4. Human Genome Sequencing Center, Baylor College of Medicine, Houston, TX
5. Marshfield Clinic Research Institute, Marshfield, WI
6. Clinical and Translational Hereditary Cancer Program, Division of Genetic Medicine, Vanderbilt-Ingram Cancer Center, Vanderbilt University, Nashville, TN
7. Department of Biomedical Informatics and Medical Education, University of Washington School of Medicine, Seattle, WA
8. School of Graduate Studies and Research, Meharry Medical College, Nashville, TN
9. Department of Microbiology, Immunology and Physiology, Meharry Medical College, Nashville, TN
10. Feinberg School of Medicine, Northwestern University, Chicago, IL
11. PRecisiOn Medicine Translational Research (PROMoTeR) Center, Department of Population Medicine, Harvard Pilgrim Health Care Institute, Boston, MA, USA, Department of Population Medicine, Harvard Medical School, Boston, MA
12. Division of Genetics, Department of Medicine, Brigham and Women's Hospital, Boston, MA,
13. McGill University Health Centre, Montreal, Quebec
14. Laboratory for Molecular Medicine, Partners Healthcare Personalized Medicine, Cambridge, MA Department of Pathology, Massachusetts General Hospital, Harvard Medical School, Boston, MA
15. Department of Biomedical informatics, Columbia University Irving Medical Center, New York, NY
16. Genetic Services and Kaiser Permanente Washington Health Research Institute, Kaiser Permanente of Washington, Seattle, WA
17. Center for Applied Genomics, Children's Hospital of Philadelphia, Philadelphia, PA
18. Division of Human Genetics, Departments of Pediatrics, The University of Pennsylvania Perelman School of Medicine, Philadelphia, PA
19. Genomic Medicine Institute, Geisinger, Danville, PA

20. Departments of Medicine (Medical Genetics) and Genome Sciences, University of Washington, Seattle, WA
21. Brigham and Women's Hospital, Broad Institute, Ariadne Labs and Harvard Medical School, Boston, MA
22. Departments of Pediatrics and Medicine, Columbia University, New York, NY
23. Division of Nephrology, Department of Medicine, Columbia University Irving Medical Center, New York, NY
24. Center for Precision Medicine and Genomics, Department of Medicine, Columbia University Irving Medical Center, New York, NY
25. Broad Institute of MIT and Harvard, Cambridge, MA
26. Medical & Population Genetics Program and Genomics Platform, Broad Institute of MIT and Harvard Cambridge, Cambridge, MA, USA. Center for Genomic Medicine, Massachusetts General Hospital, Boston, MA, USA. Department of Pathology, Harvard Medical School, Boston, MA, USA.
27. Division of Cardiovascular Medicine, Division of Clinical Pharmacology, Department of Medicine, Vanderbilt University Medical Center, Nashville, TN; Department of Pharmacology, Vanderbilt University, Nashville, TN.
28. National Human Genome Research Institute, National Institutes of Health, Bethesda, MD

**Supplemental Figures:**

**Supplemental Figure 1. Permutation test to estimate the empirical phenome-wide significant P value threshold to control type I errors.**

**Supplemental Figure 2. Number of phenotypes in the carriers of each gene in the eMERGEseq and the HCR datasets and the number of overlapped phenotypes between these two datasets.**

**Supplemental Figure 3. The PheWAS plot for *MUTYH* monoallelic carriers.**

**Supplemental Figure 4. The PheWAS plot for *APC* I1307K carriers.**

**Supplemental Figure 5. Total serum 25(OH)D levels in *BRCA1* carriers by breast cancer diagnoses.**

**Supplemental Tables**

**Table S1 Population characteristics of the eMERGEseq and HCR cohorts.**

**Table S2 Number of carriers, non-carriers and patients with a VUS variant by each hereditary cancer gene in the eMERGEseq and hereditary cancer registry studies.**

**Table S7 Conditional analyses for new associations.**

**Table S11 A scalable EHR-based approach for phenotype discovery and variant interpretation for hereditary cancer genes.**

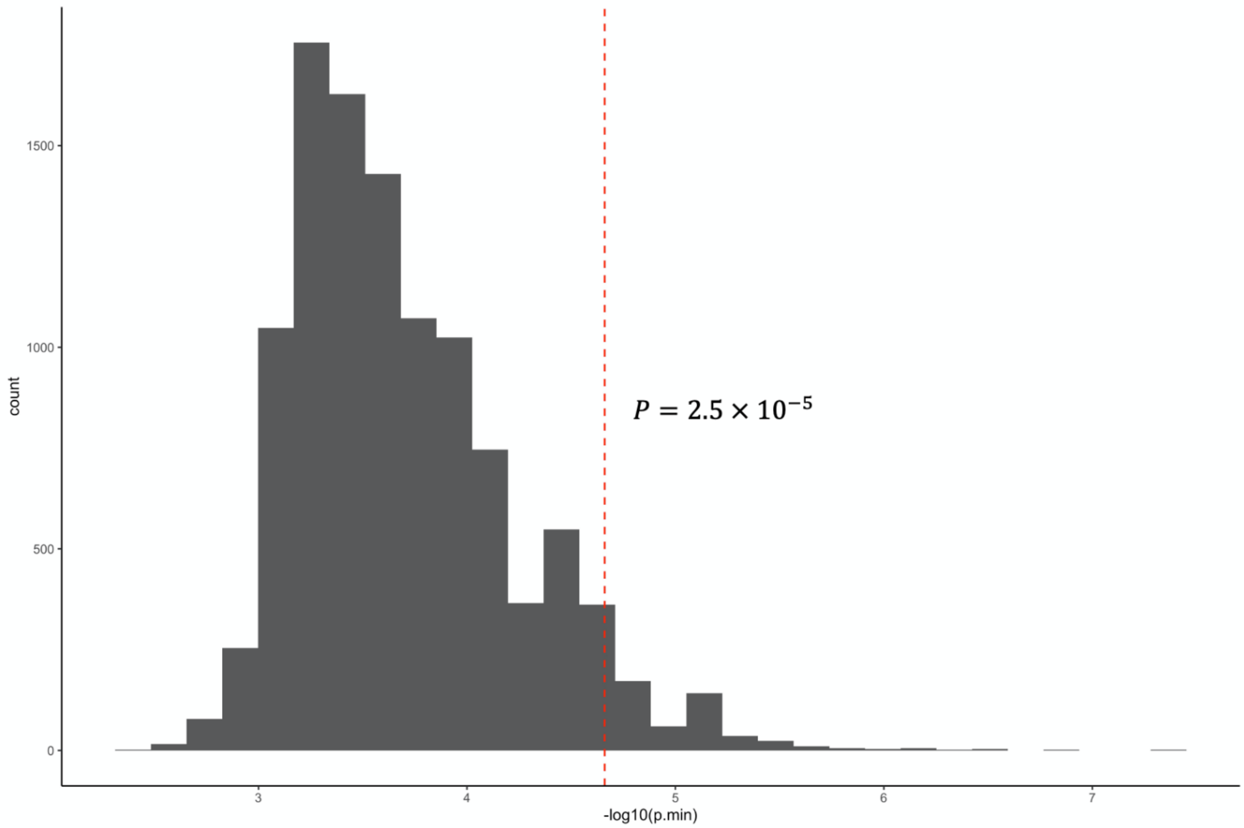


**Supplemental Figure 1. Permutation test to estimate the empirical phenome-wide significant P value threshold to control type I errors.**

We conducted 10,000 PheWASs of a random variable using firth logistic regression in the eMERGEseq data and analyzed the distributions of minimum P values (Pmin) for each PheWAS. The 95th percentile of Pmin was $2.5 \times 10^{-5}$, and we defined this P value as the empirical phenome-wide significance threshold at a significance level of $\alpha = 0.05$.

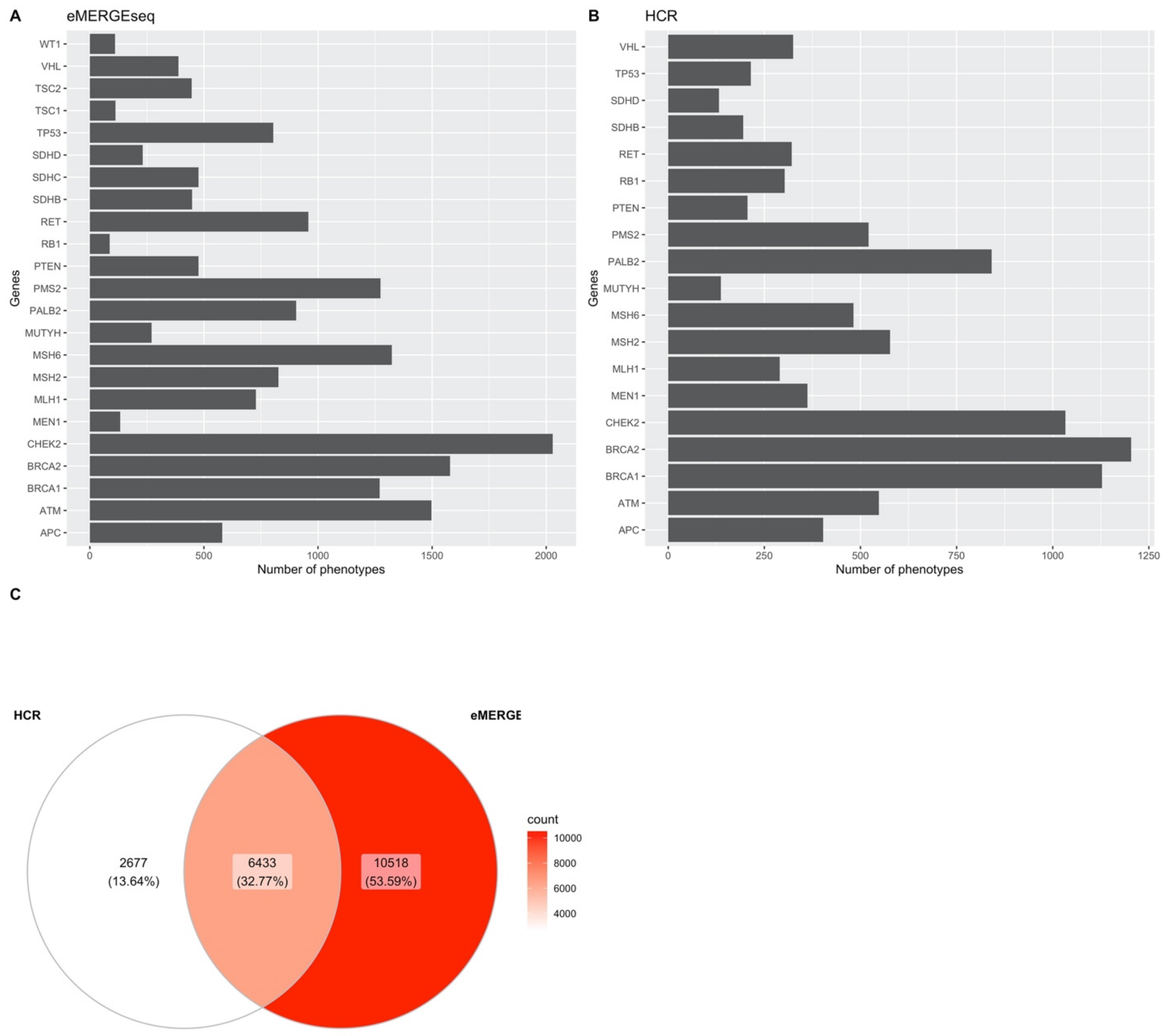


**Supplemental Figure 2. Number of phenotypes in the carriers of each gene in the eMERGEseq and the HCR datasets and the number of overlapped phenotypes between these two datasets.**

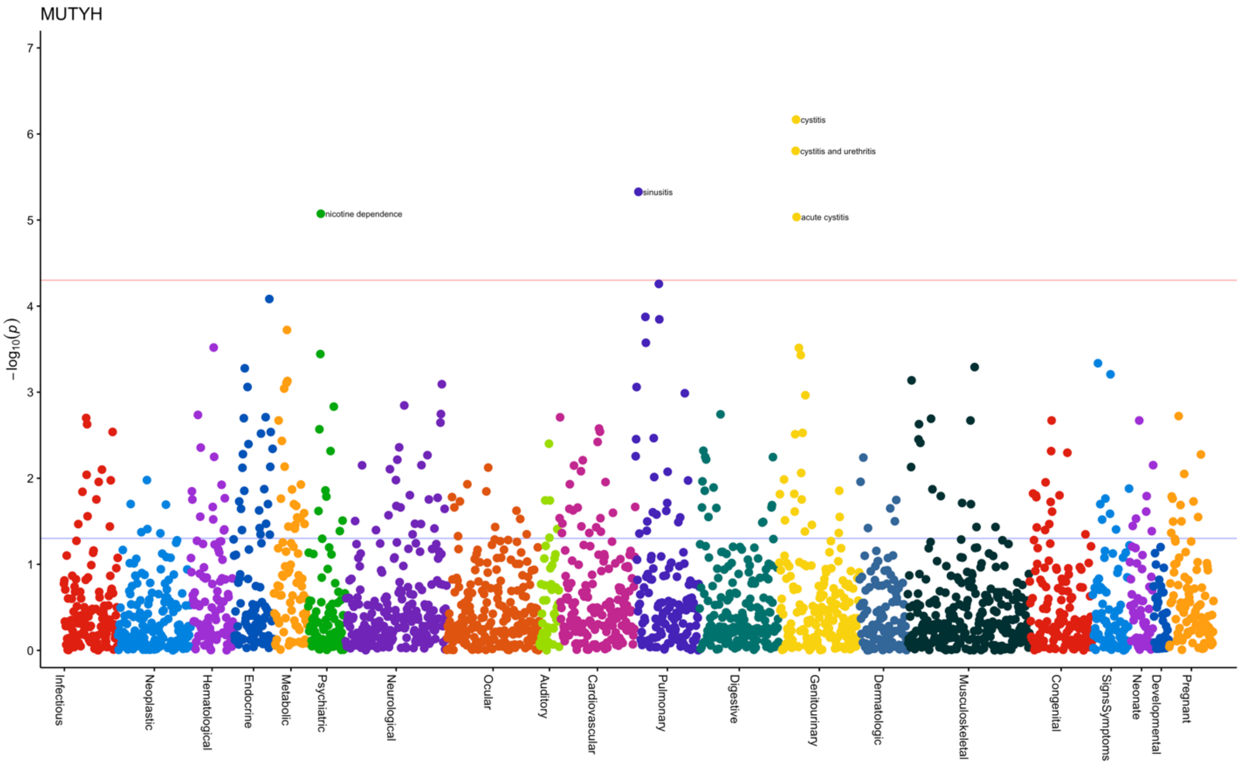


**Supplemental Figure 3. The PheWAS plot for *MUTYH* monoallelic carriers.**
The PheWAS was performed in the eMERGEseq cohort. We removed *MUTYH* monoallelic carriers from this analysis.

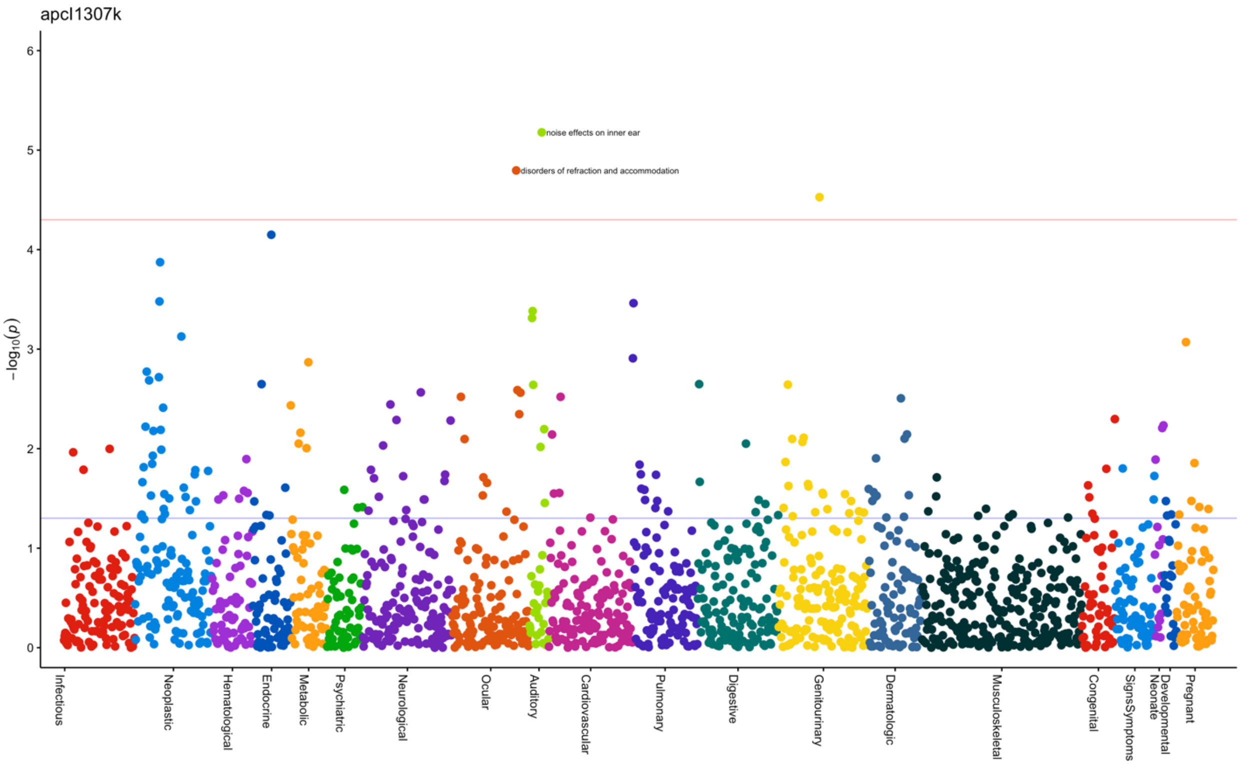


**Supplemental Figure 4. The PheWAS plot for *APC* I1307K carriers. The analysis was performed in the eMERGEseq cohort.**

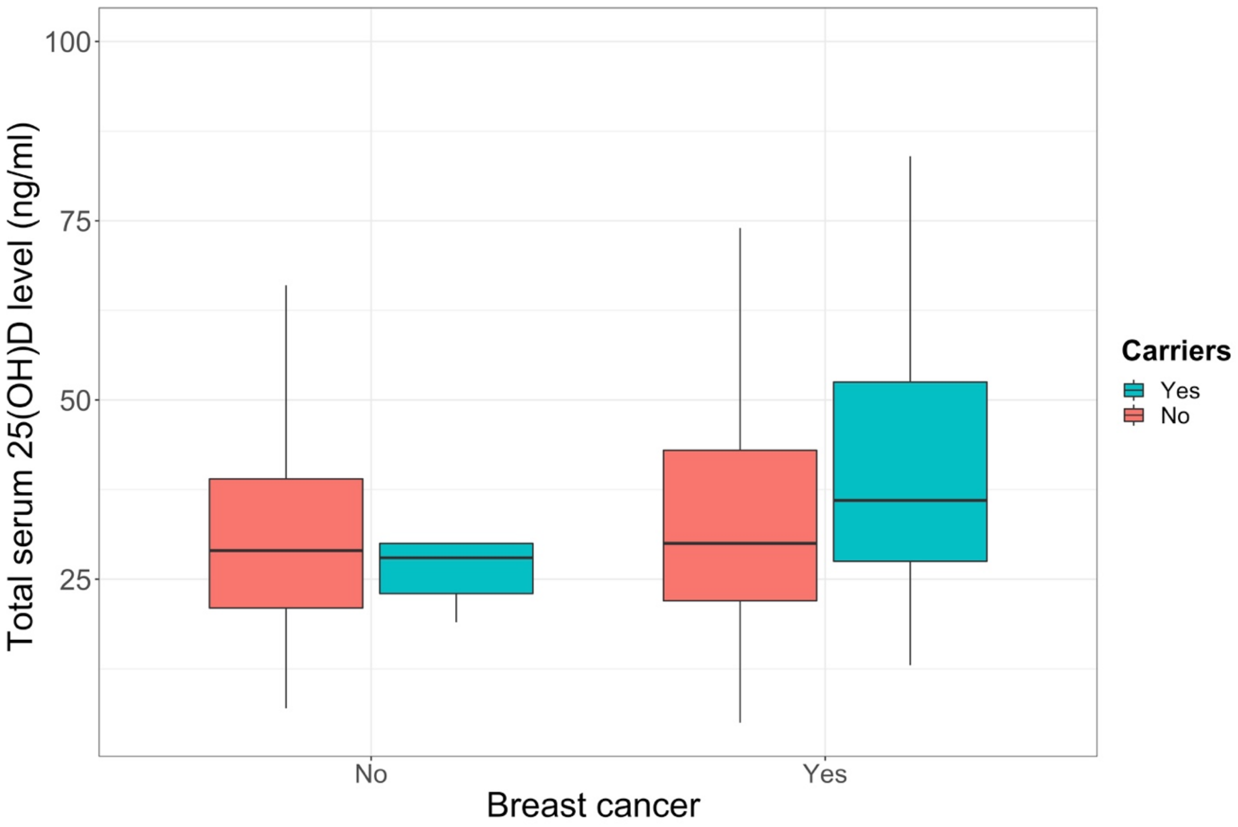


**Supplemental Figure 5. Total serum 25(OH)D levels in *BRCA1* carriers by breast cancer diagnoses**. We evaluated a total of 1,381 participants with *BRCA1* genetic testing results and data of the total serum 25(OH)D level available in the Research Derivative at Vanderbilt.

**Table S1 Population characteristics of the eMERGEseq and HCR cohorts**

| | eMERGEseq (N=23544) | | | | | | | | | HCR (N=3242) |
|---|---|---|---|---|---|---|---|---|---|---|
| **Site** | **CCHMC** | **CHOP** | **Columbia** | **Geisinger** | **Harvard** | **KPW/UW** | **Mayo** | **Northwestern** | **VUMC** | |
| **N** | 2936 | 2964 | 2314 | 2489 | 2472 | 2479 | 2439 | 2963 | 2431 | 3242 |
| **Ancestry*** | | | | | | | | | | |
| African | 1096 | 1214 | 441 | 78 | 167 | 57 | 10 | 397 | 159 | 250 |
| Asian | 32 | 83 | 190 | 19 | 75 | 919 | 17 | 146 | 18 | 21 |
| European | 1808 | 1667 | 1683 | *2392* | 2230 | 1503 | 2412 | 2420 | 2254 | 2829 |
| **Female (%)** | 1464 (50%) | 926 (31%) | 1247 (54%) | 1685 (68%) | 1404 (57%) | 1497 (60%) | 1407 (58%) | 1824 (62%) | 1133 (47%) | 2851 (88%) |
| **Age ranges (years)** | 0-80 | 0-22 | 0-89 | 0-90 | 0-91 | 32690 | 0-73 | 0-90 | 33117 | 0-99 |
| **Mean age (years)** | 16.8 | 13.9 | 52.8 | 47.8 | 57.7 | 61.6 | 62.6 | 60.5 | 66.7 | 52.5 |
| **Mean (SD) EHR follow-up (years)** | 7.3 (3.1) | 9.9 (4.0) | 8.1(7.0) | 13.1 (5.6) | 11.9 (7.4) | 18.1 (6.8) | 22.1 (6.1) | 13.0 (6.9) | 11.7 (5.4) | 8.8 (6.5) |

SD: standard deviation; CCHMC: Cincinnati Children's Hospital Medical Center; CHOP: Children's Hospital of Philadelphia; KPW/UW Kaiser Permanente Washington/University of Washington; VUMC: Vanderbilt University Medical Center. The age range for each patient was determined by the first date and the last date of ICD codes in the record. Only CCHMC, CHOP, and HCR included children.

* In the eMERGEseq cohort, the ancestry was determined by using common variants throughout the eMERGEseq panel that includes 214 ancestry informative markers, while the ancestry was self-reported in the HCR dataset.

| Table S2 Number of carriers, non-carriers and patients with a VUS variant by each hereditary cancer gene in the eMERGEseq and hereditary cancer registry studies | | | | | | | | |
|---|---|---|---|---|---|---|---|---|
| | eMERGEseq | | | | HCR | | | |
| **Gene** | **P/LP** | **VUS** | **B/LB/no rare variant** | **Percentage with P/LP** | **P/LP** | **VUS** | **B/LB/no rare variant** | **Percentage with P/LP** |
| ***APC*** | 14 | 1751 | 23191 | 0.06% | 22 | 71 | 1931 | 1.09% |
| ***APC* riskallele ( I1307K)** | 118 | 0 | 23191 | 0.51% | 1 | 0 | 1931 | 0.05% |
| ***ATM*** | 86 | 1721 | 23149 | 0.34% | 29 | 125 | 2309 | 1.18% |
| ***BLM**** | 0 | 57 | 24899 | 0 | 1 | 10 | 395 | 0.25% |
| ***BMPR1A**** | 0 | 240 | 24716 | 0 | 0 | 10 | 2133 | 0 |
| ***BRCA1*** | 86 | 481 | 24389 | 0.34% | 92 | 34 | 2705 | 3.25% |
| ***BRCA2*** | 145 | 1161 | 23650 | 0.58% | 91 | 75 | 2658 | 3.22% |
| ***CHEK2*** | 275 | 608 | 24073 | 1.10% | 45 | 50 | 2393 | 1.81% |
| ***MEN1*** | 2 | 296 | 24658 | 0.01% | 10 | 7 | 637 | 1.53% |
| ***MLH1*** | 14 | 336 | 24606 | 0.06% | 15 | 22 | 2371 | 0.62% |
| ***MSH2*** | 16 | 1245 | 23695 | 0.06% | 24 | 47 | 2347 | 0.99% |
| ***MSH6*** | 52 | 915 | 23989 | 0.21% | 16 | 44 | 2346 | 0.67% |
| ***MUTYH* biallelic** | 4 | 707 | 23788 | 0.02% | 3 | 34 | 2113 | 0.14% |
| ***MUTYH* monoallelic** | 461 | 707 | 23788 | 1.85% | 53 | 34 | 2113 | 2.41% |
| ***NF2**** | 0 | 260 | 24696 | 0 | 0 | 3 | 259 | 0 |
| ***PALB2*** | 32 | 626 | 24298 | 0.13% | 30 | 37 | 2391 | 1.22% |
| ***PMS2*** | 57 | 1307 | 23592 | 0.23% | 17 | 48 | 2326 | 0.71% |
| ***PTEN*** | 14 | 367 | 24575 | 0.06% | 3 | 21 | 2518 | 0.12% |
| ***POLD1**** | 0 | 1068 | 23888 | 0 | 0 | 18 | 1519 | 0 |
| ***POLE**** | 0 | 2431 | 22525 | 0 | 0 | 45 | 1484 | 0 |
| ***RB1*** | 2 | 472 | 24482 | 0.01% | 6 | 2 | 280 | 2.08% |
| ***RET*** | 34 | 833 | 24089 | 0.14% | 10 | 7 | 370 | 2.58% |
| ***SDHAF2**** | 0 | 90 | 24866 | 0 | 0 | 1 | 334 | 0 |
| ***SDHB*** | 6 | 155 | 24795 | 0.02% | 4 | 1 | 659 | 0.60% |
| ***SDHC*** | 6 | 207 | 24743 | 0.02% | 0 | 1 | 704 | 0.00% |
| ***SDHD*** | 4 | 110 | 24842 | 0.02% | 5 | 0 | 653 | 0.76% |
| ***SMAD4**** | 0 | 236 | 24717 | 0 | 0 | 11 | 2139 | 0 |
| ***STK11**** | 0 | 284 | 24672 | 0 | 4 | 16 | 2634 | 0.15% |
| ***TP53*** | 14 | 206 | 24736 | 0.06% | 4 | 22 | 2557 | 0.15% |
| ***TSC1*** | 5 | 770 | 24181 | 0.02% | 0 | 1 | 690 | 0.00% |
| ***TSC2*** | 13 | 2043 | 22900 | 0.05% | 0 | 11 | 680 | 0.00% |
| ***VHL*** | 5 | 228 | 24723 | 0.02% | 8 | 6 | 872 | 0.90% |
| ***WT1*** | 3 | 344 | 24609 | 0.01% | 0 | 2 | 241 | 0 |

Abbreviations: P/LP: pathogenic/likely pathogenic; B/LB: benign/likely benign; VUS: variant of uncertain significance.

*In the eMERGEseq, no P/LP variants for cancer syndromes were found for BLM, BMPR1A, NF2, POLD1, POLE, SDHAF2, SMAD4 and STK1. For SMAD4, three P/LP variants were identified for Myhre syndrome instead of cancer syndrome.

** In the HCR cohort, no P/LP variants were found SDHC, TSC1, TSC2, WT1, in addition to genes for which no P/LP variants were identified in the eMERGEseq cohort.

**Table S7 Conditional analyses for new associations**

| Gene | PheWAS phenotypes | Conditioned on | eMERGEseq | | HCR | | Meta | | |
|---|---|---|---|---|---|---|---|---|---|
| | | | OR | *P* | OR | *P* | OR | *P* | *P* heterogenicity |
| *BRCA1* | Ovarian cyst | Breast or ovarian cancer | 4.7 | 3.56E-07 | 1.3 | 0.39 | 2.3 | 3.8E-05 | 0.002 |
| *BRCA1* | Vitamin D deficiency | Breast or ovarian cancer | 0.7 | 0.12 | 0.2 | 0.0002 | 0.5 | 2.0E-02 | 0.05 |
| *BRCA2* | Ovarian cyst | Breast or ovarian cancer | 3.5 | 1.28E-06 | 2.6 | 0.0009 | 3.0 | 1.9E-09 | 0.39 |
| *APC* | Gastritis and duodenitis | Colorectal polyps or cancer | 4.9 | 0.03 | 3.7 | 0.01 | 4.1 | 6.2E-04 | 0.75 |
| *APC* | Benign neoplasm of the liver and intrahepatic bile ducts | Colorectal polyps or cancer | 53.3 | 2.37E-04 | 12.8 | 0.01 | 28.2 | 2.9E-06 | 0.24 |
| *MLH1* | Ulceration of the lower GI tract | Colorectal cancer or endometrial cancer | 7.5 | 0.009 | 11.7 | 0.01 | 10.0 | 2.8E-05 | 0.81 |
| *VHL* | Congenital malformations of spleen | VHL diagnosis or phakomatosis | 61.4 | 0.008 | 9.5 | 0.19 | 31.1 | 7.6E-04 | 0.38 |
| *PMS2* | Spermatocele | Colorectal cancer or endometrial cancer | 11.1 | 0.002 | 35.5 | 0.02 | 13.7 | 1.3E-05 | 0.46 |
| *PMS2* | Cannabis dependence | Colorectal cancer or endometrial cancer | 13.7 | 0.004 | 125.7 | 0.004 | 25.7 | 1.9E-06 | 0.08 |
| *MUTYH* | Polycystic ovarian syndrome | Colorectal polyps or cancer | 24.5 | 0.04 | 45.0 | 0.01 | 34.0 | 0.002 | 0.79 |
| *RET* | Diplopia | MEN2 diagnosis or MEN2-associated cancers: thyroid cancer and | 8.2 | 0.003 | 8.9 | 1.00 | 8.2 | 0.010 | 0.01 |
| *MEN1* | Acute pancreatitis | MEN1 diagnosis or MEN1-associated cancers including pituitary adenoma, pancreatic cancer and neuroendocrine | 3.8 | 0.39 | 4.6 | 0.21 | 4.3 | 0.17 | 0.93 |
| *CHEK2* | Leukemia | Breast cancer | 4.5 | 0.003 | 4.1 | 0.08 | 4.3 | 8.3E-05 | 0.92 |
| *MSH6* | Malignant neoplasm of the bladder | Colorectal cancer or endometrial cancer | 6.2 | 0.01 | 27.1 | 5.6E-05 | 10.4 | 4.3E-08 | 0.09 |

**Table S11. Number of carriers and differences of zPheRS between carriers and non-carriers for each gene.**

| Gene | No. of carriers | OMIM-based PheRS | | | New PheRS | | | |
|---|---|---|---|---|---|---|---|---|
| | | mean zPheRS among carriers | mean zPheRS among noncarriers | ***P*** | mean zPheRS among carriers | mean zPheRS among noncarriers | ***P*** | New phenotype incuded |
| ***APC*** | 14 | 1.3 | 0.0 | 2.00E-02 | 1.6 | 0.0 | 3.00E-02 | Benign neoplasm of the liver and intrahepatic bile ducts, gastritis and duodenitis |
| ***ATM*** | 86 | 0.2 | 0.0 | 1.00E-02 | - | - | - | - |
| ***BRCA1*** | 86 | 0.8 | 0.0 | 8.60E-19 | 1.1 | 0.0 | 3.80E-23 | Ovarian cyst |
| ***BRCA2*** | 145 | 0.6 | 0.0 | 4.30E-16 | 0.8 | 0.0 | 1.80E-22 | Ovarian cyst |
| ***CHEK2*** | 275 | 0.1 | 0.0 | 4.00E-02 | 0.1 | 0.0 | 1.90E-03 | |
| ***MLH1*** | 14 | 0.6 | 0.0 | 2.00E-02 | 0.6 | 0.0 | 2.00E-02 | Ulceration of the lower GI tract |
| ***MSH2*** | 16 | 1.6 | 0.0 | 2.20E-06 | - | - | - | - |
| ***MSH6*** | 52 | 0.9 | 0.0 | 2.90E-10 | 0.9 | 0.0 | 4.10E-10 | Premature separation of placenta, bladder cancer |
| ***PALB2*** | 32 | 0.3 | 0.0 | 1.30E-01 | - | - | - | - |
| ***PMS2*** | 57 | 0.7 | 0.0 | 3.40E-10 | 0.8 | 0.0 | 8.10E-11 | Other infection during labor, spermatocele and cannabis dependence |
| ***PTEN*** | 13 | 3.3 | 0.0 | 7.70E-07 | - | - | - | - |
| ***RET*** | 34 | 2.4 | 0.0 | 2.30E-05 | 2.5 | 0.0 | 4.60E-07 | Diplopia |
| ***TP53*** | 14 | 2.1 | 0.0 | 7.70E-06 | - | - | - | - |
| ***TSC2*** | 12 | 4.9 | 0.0 | 1.60E-09 | 5.1 | 0.0 | 1.70E-09 | Dementia |

* The P-value for ech gene were derived from the Wilcoxon rank sum test comparing the distributions of zPheRS between carriers and non-carriers.
